# Estrogen receptor negative breast cancer incidence rates are similar in Ghanaian and Non-Hispanic Black women in the USA

**DOI:** 10.1101/2022.02.21.22271266

**Authors:** Brittny C Davis Lynn, Jonine Figueroa, Fred Kwame Awittor, Naomi O. Ohene Oti, Lawrence Edusei, Nicolas Titiloye, Ernest Adjei, Beatrice Wiafe Addai, Robertson Adjei, Lucy T. Afriyie, Joel Yarney, Daniel Ansong, Seth Wiafe, Thomas Ahearn, Verna Vanderpuye, Florence Dedey, Louise A. Brinton, Baffour Awuah, Joe Nat Clegg-Lamptey, Mustapha Abubakar, Montserrat Garcia-Closas, Richard Biritwum, Barry I Graubard, the Ghana Breast Health Study Team

**Affiliations:** Division of Cancer Epidemiology and Genetics, National Cancer Institute; Usher Institute, Institute of Genetics and Cancer, University of Edinburgh, Edinburgh, Scotland; Komfo Anokye Teaching Hospital, Kumasi, Ghana; University of Ghana, Accra, Ghana; Korle Bu Teaching Hospital, Accra, Ghana; Peace and Love Hospital, Kumasi, Ghana; Komfo Anokye Teaching Hospital, Kumasi, Ghana; Ghana Statistical Service, Accra, Ghana; Peace and Love Hospital, Kumasi, Ghana; Kwame Nkrumah University of Science and Technology, Kumasi, Ghana; Ghana Statistical Service, Accra, Ghana; Loma Linda University, Loma Linda, CA; Kwame Nkrumah University of Science and Technology, Kumasi, Ghana; University of Ghana, Accra, Ghana; Loma Linda University, Loma Linda, CA

**Author notes:** Contributed equally (Brittny C Davis Lynn, Jonine Figueroa, Fred Kwame Awittor, Naomi O. Ohene Oti, Montserrat Garcia-Closas, Richard Biritwum, Barry I Graubard). Corresponding Authors Montserrat Garcia-Closas, National Cancer Institute, Division of Cancer Epidemiology & Genetics, Office of the Director, NCI Shady Grove, Jonine Figueroa, Usher Institute, University of Edinburgh, Teviot Place, Room 3.736A. **NOTES:**. **Role of the funder:** The funder had no role in the writing of this editorial or the decision to submit it for publication. **Disclosures:** The authors have no disclosures. **Author contributions:** Writing, original draft—BCD, JDF, BIG, RB, MGC. Writing, editing and revision—All authors. Data/sample collection- LE, NT, EA, BWA, JY, FKA, NO, DA, SW, VV, FD, BA. Population control selection--RA, LTA RB. Data interpretation—All authors.

**Keywords:** breast neoplasm, age-specific incidence, disparities, estrogen receptor

## Abstract

Age-standardized incidence rates of estrogen receptor negative (ERN) breast cancers in the US are higher among Non-Hispanic Black (NHB) compared to Non-Hispanic White (NHW) women. We aimed to determine if incidence rates were similar between NHB and Ghanaians, given that a high proportion of NHB share West African genetic ancestry. We compared US rates (per 100,000 women) to those in Ghana, using data from US SEER, the Ghanaian census, and the Ghana Breast Health Study (GBHS), a population-based case-control study conducted between 2013-2015 in Accra and Kumasi. ERN age-standardized rates were similar among Ghanaian (40.7) and US NHB women (43.1), and both were higher compared to US NHW (24.0). Estrogen receptor-positive (ERP) rates were lower in Ghanaian (43.7) than US NHB (84.4), and highest in US NHW (128.5). Our data support higher ERN rates among women in Ghana similar to US NHB suggesting shared putative risk factors that require investigation.

---

In the United States (US), Non-Hispanic Black (NHB) women have higher incidence rates of estrogen receptor negative (ERN) breast tumors compared to Non-Hispanic White (NHW) women after accounting for potential confounders [1, 2]. One hypothesis for these observations is that women of African ancestry could be genetically predisposed to ERN breast cancer [3]. This hypothesis is supported by the observation of a high frequency (∼50%) of ERN breast cancers [4] in black women in sub-Saharan African (SSA), who have shared ancestry with US NHB (∼70% of African-Americans have SSA ancestry), particularly in West Africa [5]. However, ERN tumors are frequently diagnosed at younger ages hence, the higher ERN frequency could just reflect the younger demographics in SSA countries compared to the US.

Age-standardized incidence rates rather than proportions are required to make comparisons across countries with different demographics. These comparisons have been limited by lack of high-quality data on ER-specific incidence rates in SSA countries due to study design issues such as unrepresentative case series and unstandardized tissue processing or ER staining [4]. In this report, we estimated age-specific ERN breast cancer incidence rates in Ghana and compared them to rates in NHW US to evaluate if women from Ghana (which share some genetic ancestry with NHB) and the US have similar ERN rates. We further compared ERN to ERP rates for breast cancer in these populations. We used data from the Ghana Breast Health Study (GBHS), a population-based case-control study conducted during 2013-2015 in two major cities in Ghana, Accra and Kumasi, and census data to generate sampling fractions and estimate crude, age-specific and standardized incidence rates of breast cancer overall and by ER status (see **Supplemental Methods**) [6, 7]. Rates for NHW and NHB US women during 2013-2015 were based on data from 17 US SEER registries [8].

Age and tumor characteristics for women diagnosed with breast cancer in GBHS and SEER are presented in **Table 1**. Women in Ghana are diagnosed at earlier ages and have a higher frequency of ERN tumors compared to the US. However, the age-standardized rates for ERN tumors are similar for Ghanaian (40.7 per 100,000) and NHB (43.1) women, while NHW women had substantially lower rates (24.0). In contrast, ERP age-standardized rates are substantially lower in Ghanaian (43.7) than either US NHB (105.4) or NHW (128.5) women. Age-specific incidence rates for breast cancer overall and by ER status for NHW and NHB women, aged 20-74 years, are presented in **Figure 1**. While the overall age-specific incidence rates in Ghana were lower than in the US, they show a similar trend of rapid rise between 20 to 50 years followed by a slowing of rates around 50, the approximate age at menopause. Age-specific breast cancer rates by ER status in Ghana (**Figure 1C**) show rates are similar by ER status, with some suggestion of cross over about age 35. This pattern is different from NHB and NHW women in the US, where ERP rates are higher than ERN rates across all age groups (**Supplemental Figure 1**). This is consistent with previous analyses of US SEER data for breast cancer cases diagnosed from 2013-2015 [9]. While ERP rates are higher in US NHB than in Ghanaian women, ERN rates appear to be similar in the two populations (**Figure 1 B-C)**, as shown by the age-standardized rates in **Table 1**.

**Table 1:** Age-standardized rates using the World Standard (Segi, 1960) per 100,000) and incidence rate ratios (IRR) of breast cancer cases in women from Ghana (GBHS) compared to Non-Hispanic White (NHW) and Non-Hispanic Black (NHB) women in the USA (17 SEER Registries), during 2013-2015.

| Characteristic | NHW, SEER 17 |  | NHB, SEER 17 |  | GBHS |  |
| --- | --- | --- | --- | --- | --- | --- |
| Age range (years) | 20-74 |  | 20-74 |  | 18-74 |  |
| Number of cases | 103,227 |  | 18,321 |  | 1,071 |  |
| *Variable* | *N* | *%* | *N* | *%* | *N* | *%* |
| Age at diagnosis (years) |  |  |  |  |  |  |
| <40 | 4459 | 4% | 1359 | 7% | 221 | 21% |
| 40-49 | 15931 | 15% | 3506 | 19% | 316 | 30% |
| 50-59 | 29063 | 28% | 5820 | 32% | 281 | 26% |
| >60 | 53774 | 52% | 7636 | 42% | 253 | 24% |
| ER expression |  |  |  |  |  |  |
| Positive (ERP) | 85,432 | 85% | 12,562 | 71% | 390 | 51% |
| Negative (ERN) | 15,040 | 15% | 5,117 | 28% | 380 | 49% |
| Unknown | 2,755 |  | 642 |  | 301 |  |
|  | Rate | 95%CI | Rate | 95%CI | Rate | 95%CI |
| Overall crude incidence rate | 205.8 | (204.6-207.1) | 165.6 | (163.2-168.0) | 59.2 | (55.7-62.8) |
| Overall age-standardized rate* | 152.9 | (151.9-153.8) | 148.5 | (146.4-150.7) | 84.4 | (79.2-89.9) |
| ERP crude incidence rate | 175.0 | (173.8-176.2) | 117.7 | (115.7-119.7) | 29.9 | (27.5-32.6) |
| ERP age-standardized rate* | 128.5 | (127.9-129.7) | 105.4 | (103.6-107.3) | 43.7 | (40.0-47.8) |
| ERN crude incidence rate | 30.8 | (30.3-31.3) | 47.9 | (46.7-49.2) | 29.2 | (26.9-31.9) |
| ERN age-standardized rate* | 24.0 | (23.6-24.4) | 43.1 | (42.0-44.3) | 40.7 | (37.2-44.5) |

\*ER expression % are excluding Unknown status.

**Figure 1:**
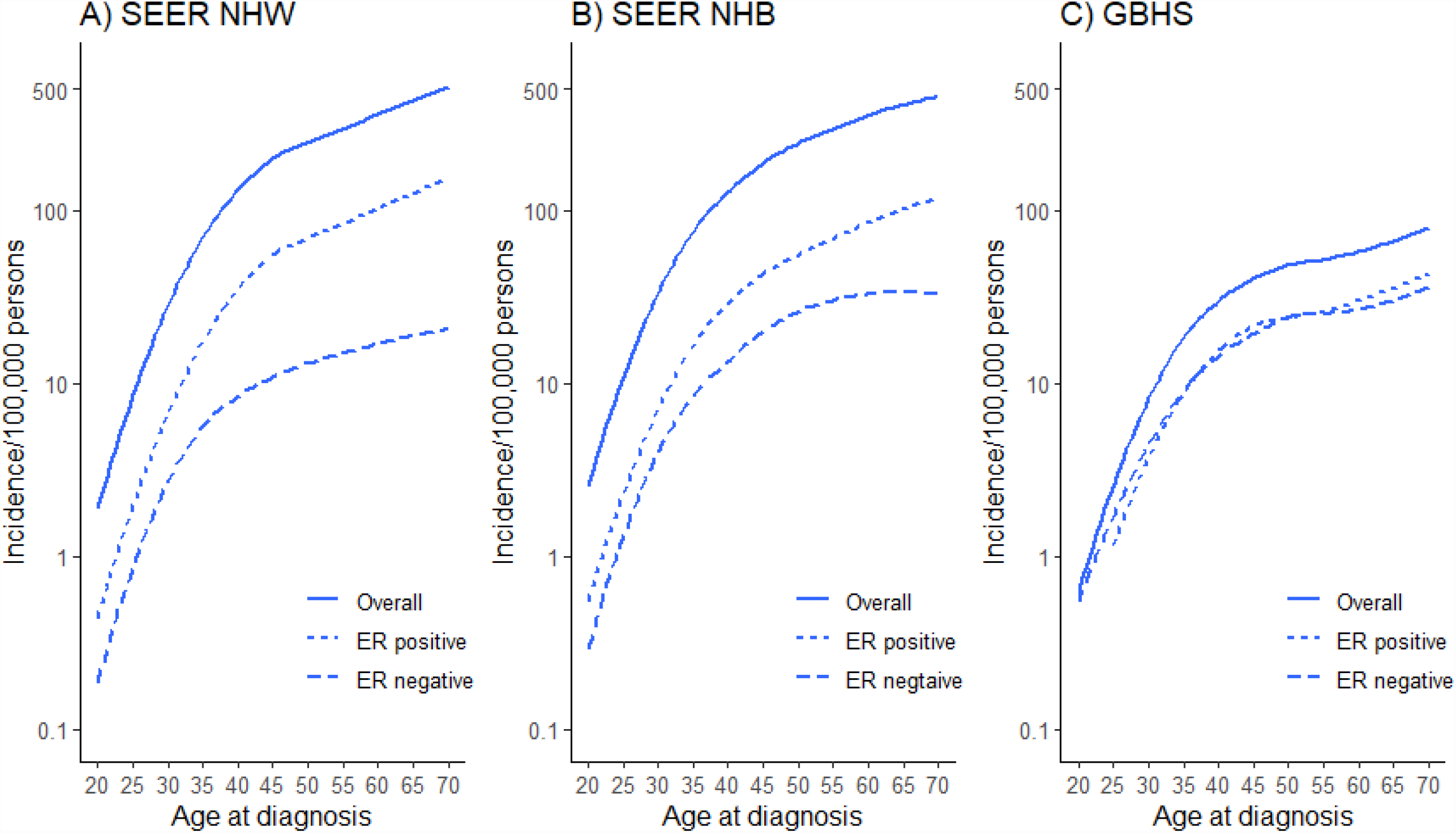
Age-specific incidence by ER status for NHW and NHB women from 17 SEER registries and for women from the GBHS. Age-standardized rates were estimated using the World Standard (Segi, 1960) per 100,000), which is used in IARC Globocan data, to facilitate international comparisons. ER expression % are excluding Unknown status.

Our results are consistent with women from West Africa being genetically predisposed to developing ERN breast cancer. This is supported by higher frequencies of *BRCA1* and *BRCA2* pathogenic mutations in Ghana (Ahearn et al., submitted) and Nigeria [10, 11] compared to European-ancestry populations [12], including African Americans diagnosed with breast cancer <35 years of age [13], but in the recent carriers study, there was insufficient evidence [14]. Polygenic risk could also contribute to higher absolute risks of ERN breast cancer in women of African ancestry [15]. Consistent with previous reports [16, 17], the GBHS showed that parity is a risk factor for early-onset ERN cancers [17], while breastfeeding is inversely related to ERN risk [16, 17]. Given that both parity and breastfeeding are higher in Ghana compared to the US, it is unclear if these factors could contribute to differences in ERN rates across countries [17]. Whether biological, behavioural or social/environmental factors contribute to higher incidence rates of ERN tumours in women of SSA origins requires further investigation [1, 18], especially since data support different distributions of breast cancer subtypes in Kenya [19], South Africa [20] compared to Ghana [17]. In contrast, ERP breast cancer incidence remains low in Ghana compared to US NHB or NHWs. However, this could change as breast cancer incidence rates are rapidly rising in SSA countries [21-23] likely due to longer life expectancies, and adoption of Western lifestyle associated with risk of ERP breast cancer (e.g. obesity, later ages of first birth and fewer number of children) [17, 24]. The lack of population mammography screening in Ghana is also likely to contribute to differences in ERP rates with the US [25].

Strengths of the GBHS are the large size and population-based design in the two largest cities of Ghana, and the standardized collection and quality immunohistochemical data [17]. The estimated age-standardized rate for overall breast cancer in Ghana (84.4 cases per 100,000 (95% CI 79.2-89.9)) is higher than the 62.4 cases per 100,000 reported by Globocan [26]. This could be due to higher rates in urban areas in the GBHS compared to rural areas that could result in an underestimation of observed rate differences with the US. Another limitation was missing data on ER status (28% of cases) that was addressed by multiple imputation of missing data [27].

The similar rates observed for ERN tumours in US NHB and Ghanaians supports the hypothesis of genetic predisposition for ERN breast cancer particularly for West African ancestry women, further population-based studies are needed to delineate biological, behavioral, and social factors contributing to the higher ERN rates in SSA where mortality rates remain the highest globally [26].

## Data Availability

All data produced in the present study are available upon reasonable request to the authors

## Acknowledgements

This research was supported by the Intramural Research Program and the Cancer Prevention Fellowship Program of the National Cancer Institute at the National Institutes of Health.

