## Supplemental material for "Estrogen receptor negative breast cancer incidence rates are similar in Ghanaian and Non-Hispanic Black women in the USA"

**Supplemental Methods**

The GBHS has been previously described [6, 17]. Briefly, breast cancer cases were recruited from three hospitals in Accra and Kumasi, including Peace and Love Hospital, Korle Bu Teaching Hospital, and Komfo Anoyke Teaching Hospital. Population controls (n=2,106) were frequency matched by age group. Enumeration efforts for control selection was performed using systematic random sampling and have been described previously [6, 7, 17].

Sample weights for cases and controls were calculated separately. Analysis included invasive breast cancer case that were histologically confirmed (n=1,071, 95% of 1,126 [17]) with known age in the GBHS was given a weight of “1” as all cases were used in this study. Weights for controls were calculated by district and 5-year age group (18-24, 25-29, 30-34, 35-39, 40-44, 45-49, 50-54, 55-59, 60-64, 65-69, and 70-74 years) at enumeration. A sample fraction for each age group by district was derived from the sample design. The sample fraction was adjusted by the nonresponse rate and then a poststratification adjustment was used so that the sample weights (i.e., the inverse of these adjusted sample fractions) summed over the controls selected from each district and age group matched the 2010 Ghana Census population total.

Invasive breast cancer incidence data for breast cancer overall as well as by ER status were obtained from 17 Surveillance, Epidemiology, and End Results (SEER) Program registries for the years 2013-2015 for non-Hispanic Black (NHB) and non-Hispanic White (NHW) women aged 20-74 years.

Sequential regression algorithm multiple imputation, which addresses missing data in the independent variables, was performed by using series of logistic regressions to predict ER status for cases from age, site, grade of tumor, age of menarche, and number of pregnancies. Breast cancer in GBHS tended to be younger and had a higher percentage of unknown ER status compared to SEER (Supplemental Table 1). ER Imputation was completed separately for GBHS data as well as for the SEER data by race/ethnicity [27, 28].

To calculate age-specific incidence rates of breast cancer in Ghana from the GBHS data, weights were applied to the controls to estimate the overall population of women in Ghana. Control weights were generated by district and age group using enumeration data from the 2010 Census in Ghana. Age-specific incidence rates were calculated by taking the total number of invasive breast cancer cases from the GBHS and dividing by the weighted population-based controls. This ratio was divided by 3 years and 100,000 persons to report the 3-year rate per 100,000 women in Ghana. These rates were calculated for each 5-year age group overall and by risk factors. The logarithm of age-specific incidence rates for each risk factor were plotted. Wald test was used to test for interaction between age-specific rates by risk factors. Age specific incidence by ER status was plotted using the population level data with the imputations.

Supplemental Figure 1 Age-specific percentage breast cancer in SEER and Ghana by estrogen receptor (ER) status

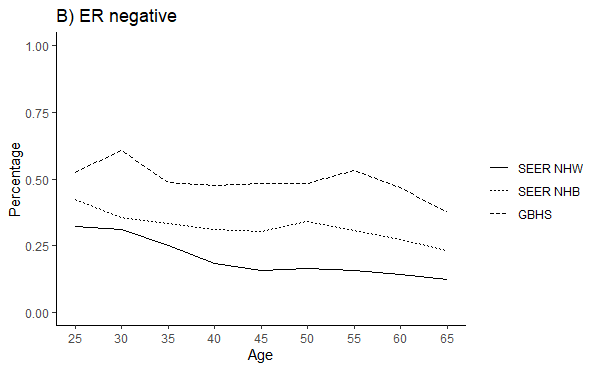

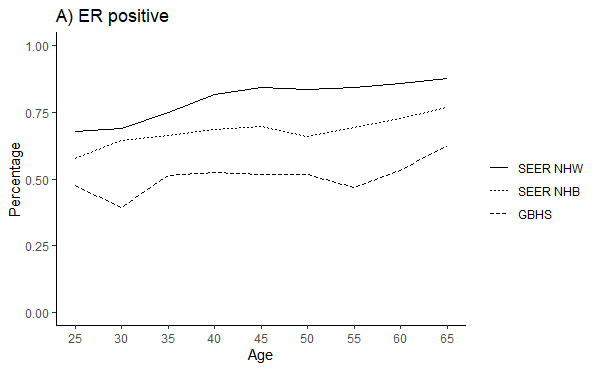

USA SEER and Ghana breast cancer invasive breast cancers diagnosed from 2013-2015 by ER status. NHW (Non-Hispanic Whites), NHB (Non-Hispanic Blacks)

Supplemental table 1: Frequency distribution of tumor characteristics and factors used in for multiple imputation in the Ghana Breast Health Study

|  | **ER positive (n=390)** | | **ER Negative (n=380)** | | **ER Missing (n=301)** | |
| --- | --- | --- | --- | --- | --- | --- |
| variable | n | % | n | % | n | % |
| **Age** |  |  |  |  |  |  |
| <35 | 29 | 7% | 44 | 12% | 34 | 11% |
| 35-44 | 94 | 24% | 87 | 23% | 84 | 28% |
| 45-54 | 121 | 31% | 113 | 30% | 78 | 26% |
| 55+ | 146 | 37% | 136 | 36% | 105 | 35% |
| **Tumor Grade** |  |  |  |  |  |  |
| 1 | 21 | 5% | 10 | 3% | 10 | 3% |
| 2 | 107 | 27% | 52 | 14% | 44 | 15% |
| 3 | 189 | 48% | 241 | 63% | 128 | 43% |
| Unknown | 73 | 19% | 77 | 20% | 119 | 40% |
| **Tumor Size** |  |  |  |  |  |  |
| <=2cm | 14 | 4% | 14 | 4% | 10 | 3% |
| >2-5cm | 112 | 29% | 118 | 30% | 96 | 25% |
| >5cm | 228 | 58% | 214 | 55% | 138 | 35% |
| No palpable lump | 2 | 1% | 0 | 0% | 1 | 0% |
| small/medium | 0 | 0% | 0 | 0% | 1 | 0% |
| Unknown | 34 | 9% | 44 | 11% | 144 | 37% |
| **Site** |  |  |  |  |  |  |
| KATH | 84 | 22% | 115 | 30% | 80 | 27% |
| KBTH | 135 | 35% | 119 | 31% | 120 | 40% |
| PLH | 171 | 44% | 146 | 38% | 101 | 34% |
| **Education** |  |  |  |  |  |  |
| No formal education | 76 | 19% | 92 | 24% | 70 | 23% |
| Primary school | 62 | 16% | 51 | 13% | 33 | 11% |
| Junior secondary school | 77 | 20% | 99 | 26% | 69 | 23% |
| ≥ Senior secondary school | 150 | 38% | 110 | 29% | 115 | 38% |
| Unknown | 25 | 6% | 28 | 7% | 14 | 5% |
| **Family history of breast cancer** |  |  |  |  |  |  |
| No | 361 | 93% | 349 | 92% | 275 | 91% |
| Yes | 24 | 6% | 26 | 7% | 22 | 7% |
| Unknown | 5 | 1% | 5 | 1% | 4 | 1% |
| **Body size** |  |  |  |  |  |  |
| Slight | 85 | 22% | 78 | 21% | 80 | 27% |
| Average | 144 | 37% | 141 | 37% | 128 | 43% |
| Slightly heavy | 95 | 24% | 95 | 25% | 59 | 20% |
| Heavy | 41 | 11% | 39 | 10% | 19 | 6% |
| Unknown | 25 | 6% | 27 | 7% | 15 | 5% |
| **Age at menarche (years)** |  |  |  |  |  |  |
| < 15 | 91 | 23% | 91 | 24% | 77 | 26% |
| 15 | 94 | 24% | 82 | 22% | 63 | 21% |
| 16 | 74 | 19% | 67 | 18% | 72 | 24% |
| ≥ 17 | 89 | 23% | 80 | 21% | 46 | 15% |
| Unknown | 42 | 11% | 60 | 16% | 43 | 14% |
| **Parity** |  |  |  |  |  |  |
| Nulliparous | 35 | 9% | 33 | 9% | 33 | 11% |
| 1-2 | 110 | 28% | 98 | 26% | 94 | 31% |
| 3-4 | 136 | 35% | 118 | 31% | 95 | 32% |
| ≥ 5 | 109 | 28% | 131 | 34% | 75 | 25% |
| Unknown | 0 | 0% | 0 | 0% | 4 | 1% |
| **Age at first birth** |  |  |  |  |  |  |
| <20 | 77 | 20% | 89 | 23% | 63 | 21% |
| 20-25 | 94 | 24% | 94 | 25% | 65 | 22% |
| 25-30 | 85 | 22% | 92 | 24% | 67 | 22% |
| 30+ | 77 | 20% | 54 | 14% | 59 | 20% |
| unknown | 57 | 15% | 51 | 13% | 47 | 16% |
| **Ever Breastfed** |  |  |  |  |  |  |
| Never | 45 | 12% | 43 | 11% | 38 | 13% |
| Ever | 323 | 83% | 311 | 82% | 251 | 83% |
| Unknown | 22 | 6% | 26 | 7% | 12 | 4% |
| **Menopausal Status** |  |  |  |  |  |  |
| Premenopausal | 168 | 43% | 160 | 42% | 140 | 47% |
| Postmenopausal | 220 | 56% | 220 | 58% | 161 | 53% |
| Unknown | 2 | 1% | 0 | 0% | 0 | 0% |
